# Risk of Inflammatory Bowel Disease Following Hospitalisation with Infectious Mononucleosis: A Danish Nationwide Cohort Study (1977-2021)

**DOI:** 10.1101/2024.03.25.24304776

**Authors:** Anthony Ebert, Shania Harper, Marie V. Vestergaard, Wayne Mitchell, Tine Jess, Rahma Elmahdi

## Abstract

**Background:** Infectious mononucleosis (IM) is a largely self-limiting syndrome mainly affecting adolescents and young adults but can present as a more severe disease requiring hospitalisation. The infectious agent most commonly causing IM, Epstein-Barr virus (EBV) has been associated with the development of several immune-mediated and inflammatory diseases.

**Objective:** To investigate the risk of inflammatory bowel disease (IBD) following hospitalisation with severe IM.

**Methods:** Danish nationwide registries were used to identify severe IM patients and sex-, age- and municipality-matched non-IM hospitalised controls, from 1st January 1977 to 31st December 2022. We undertook Cox regression modelling to calculate the hazards (HR) and 95% CI of IBD diagnosis, including Crohn’s disease (CD) and ulcerative colitis (UC). Analyses were stratified by sex and age at IM hospitalisation.

**Results:** We identified 39,684 patients with severe IM who were sex-, age-, and municipality-matched to 396,840 non-IM hospitalised controls. Severe IM was significantly associated with the development of IBD (HR:1.35; 95% CI: 1.22-1.49) and this was seen particularly in CD (HR: 1.56; 95% CI: 1.34-1.83) and to a lesser extent in UC (HR: 1.23; 95% CI: 1.08-1.40). Sex at severe IM diagnosis was not found to be a significant modifier to the risk of IBD development with risk increased in both females (HR: 1.36; 95% CI: 1.20-1.55) and males (HR: 1.34; 95% CI: 1.17-1.54). Only those receiving a severe IM diagnosis at 10-16 years (HR: 1.42; 95% CI:1.22-1.64) or 17-29 years (HR: 1.34; 95% CI:1.15-1.56) were at increased risk of IBD development.

**Conclusion:** This study demonstrated an association between IM hospitalisation and later IBD development, indicating an association between severe EBV disease and IBD development. Further exploration of factors contributing to IBD susceptibility following EBV infection is warranted.

## INTRODUCTION

Inflammatory bowel disease (IBD) commonly diagnosed as either Crohn’s disease (CD) or ulcerative colitis (UC) is a progressive immune-mediated inflammatory disorder (IMID) of the gastrointestinal tract, affecting >6.8 million individuals worldwide.^1^ IBD is most prevalent in high-income countries whilst its incidence is most rapidly increasing in newly industrialised countries.^2^ Approximately 0.5% of the Danish population had an IBD diagnosis in 2010 and by 2020, this rose to almost 0.75% as IBD prevalence increased by 63.3%,^3^ supporting the projection that 1% of individuals within high-income countries will have an IBD diagnosis by 2030.^1^ The pathophysiology of IBD involves complex genetic, environmental, epithelial, microbial, and immune factors^4^ that result in immune dysfunction leading to gastrointestinal barrier dysregulation, and gut dysbiosis.^5^ Environmental exposures such as smoking, pro-inflammatory food and antibiotic use do not fully account for why IBD develops in genetically susceptible individuals.^6,7^ Infectious agents as either contributing to or protecting from the development of IBD have largely been associated with disease in the context of GI specific pathogens, including *Salmonella* and *Campylobacter* infection,^8–10^ or *Helicobacter pylori* infection in the induction of immune tolerance and reduced risk of IBD.^11,12^ The association between systemic viral infections and IBD development has yet to be systematically explored.

Infectious mononucleosis (IM) is a clinical syndrome of fever, tonsillar pharyngitis, and lymphadenopathy, commonly (>90% of cases) due to acute infection with Epstein-Barr Virus (EBV).^13,14^ Although usually a benign, self-limiting condition, in approximately 5% of acute infection, IM is associated with severe disease presenting with by high fever, mesenteric adenitis, and extreme fatigue, which can require hospitalisation.^15,16^ Associations between IM and several IMIDs including Sjögren’s syndrome, rheumatoid arthritis, systemic lupus erythematosus^17–19^ and multiple sclerosis (MS)^20,21^ have previously been identified and the putative role of EBV in MS development has been shown most recently in a longitudinal analysis of a cohort of more than 10 million American military personnel, undertaken by Bjornevik et al., in 2022.^22^

Preliminary evidence from a study using a bacteriophage immunoprecipitation sequencing technique, like that used by Bjornevik et al., 2022, on a cohort of 119 military service personnel diagnosed with IBD and healthy matched controls, found that exposure to EBV was significantly associated with CD, supporting a biological association with EBV exposure and IBD development.^23^ More recently, Loosen et al., 2023,^24^ undertook a retrospective cohort study using data from a subset of German primary care physicians in the Disease Analyzer Database (IQVIA) to explore the association between general practitioner recorded IBD following IM diagnosis. Although authors observe a significant association with between IM and subsequent IBD, several limitations in study design, including short follow-up time, small cohort size and lack of population representative data (only from 3% of German primary care physicians were included in the analysis), and particularly a lack of clinical data on IM severity and presentation, and validity of IBD diagnoses make interpretation of these study findings challenging.

Therefore, to explore the association between severe infectious mononucleosis (IM) and the incidence of IBD, we undertook a nationwide, population-representative cohort study using data from the Danish National Health registries. The primary outcome was the incidence of IBD, including CD and UC following hospitalisation with IM. We additionally explored whether previous hospitalisation with IM impacted on the severity of disease in those going on to develop IBD.

## MATERIALS & METHODS

### Data Sources

We used the Danish National Civil Register (CPR) to identify all individuals living in Denmark between 1st January 1977 and 31st December 2021 and linked these data at an individual level, using a unique personal identification number assigned to all Danish residents, to the Danish National Patient Register (LPR) and the Danish National Prescription Register (DPR). Through the CPR, information such as date of birth, sex, municipality of residence and date of immigration or emigration is available. All hospital admissions with diagnosis codes have been recorded in the LPR since 1977. The DPR contains individual-level information, including Anatomical Therapeutic Chemical (ATC) classification code, dose and pack size for all prescriptions redeemed at outpatient pharmacies in Denmark since 1994. As a CPR number is required for all diagnoses recorded in the LPR and prescriptions redeemed in the DPR, data from these registries are not only representative of the entire resident Danish population but are additionally complete, i.e., these data comprise all national health system hospital diagnoses or publicly reimbursed prescription in Denmark.

### Study Population

We identified individuals hospitalised with infectious mononucleosis from 1st January 1977 to 31st December 2021 to create the severe IM group. We matched these index cases at a ratio of 1:10 to any individual in the CPR without a record of hospitalisation with IM in the LPR based on year of and sex (male or female) at birth and municipality of residence. The case and their matched controls must be resident in Denmark for at least 9 years in the last 10 years to ensure incident cases are identified. The exposure, severe IM, was defined as any hospital diagnosis with infectious mononucleosis (ICD-8 codes 07500, 07501, 07508 and 07509 or ICD-10 codes B27.0, B27.1, B27.8 and B27.9) in the LPR. We further classified severe IM exposure by the presence of ICD-10 coded IM complications including hepato-splenomegaly (R16), lymphadenitis (R59, L04 or I88) or splenic rupture (D73.5; see Supplementary Table 1 for complete list of diagnostic codes) using the LPR.

The primary outcome was diagnosis with IBD, including CD or UC (ICD-8 codes 56308–09 and ICD-10 code K50 for CD; ICD-8 codes 56319 and 56904 and ICD-10 code K51 for UC). We restricted the outcome of IBD to those with two or more recorded in-patient or out-patient hospital contacts within two years, with the last recorded subtype taken as the classification for CD or UC to ensure validity of the IBD outcome.

We also assessed whether a history of severe-IM is associated with severe IBD within IBD patients using a a composite variable of IBD-related hospitalisation, surgery, or administration of biological therapies or systemic steroids (see Supplementary Table 2 for diagnostic codes for composite IBD severity score) in the LPR or DPR. Since these measures are based on ICD-10 codes, this analysis is restricted to patients diagnosed with IBD after 1994.

### Statistical Analysis

We undertook Cox proportional regression analysis to calculate the hazard ratio (HR) and 95% confidence interval (95% CI) of IBD diagnosis in those with severe IM compared their sex-, age- and municipality-matched non-severe IM counterparts by following the cohort from the index date to the date of second diagnosis with IBD. Individuals were censored from analysis at the point of death, emigration, or end of study period (31st December 2021), whichever came first. Analysis was also undertaken for risk of IBD diagnosis in severe IM compared with non-severe IM for CD and UC separately. We stratified analysis by sex, age at IM-hospitalisation (0-9 years, 10-16 years, 17-29 years, 30-59 years, and >60 years), calendar year of IM hospitalisation (1977-1989, 1990-1999, 2000-2009 and, 2010-2021), and duration of IM-hospitalisation (0-7 days, 8-14 days and >15 days). Association between severe IM and risk of severe IBD development was also assessed using Cox regression analysis by following those diagnosed with IBD for the first event from of a composite variable for IBD disease severity in the severe IM group compared with the non-severe IM group. These analyses were also stratified by sex, age at severe IM diagnosis, calendar year of diagnosis, and duration of hospitalisation.

### Sensitivity Analysis

To assess the impact of unmeasured and systemic confounding, such as positive health seeking behaviours, on the association between severe IM and IBD, we undertook negative control matching analysis exploring whether chlamydia trachomatis, an infection with an age-distribution like that of infectious mononucleosis, was also associated with an increased risk of IBD development. We undertook survival analysis to assess hazards of IBD diagnosis following infection with severe IM compared to infection with chlamydia (see Supplementary Table 1 for complete list of diagnostic codes).

Finally, we used the Danish nationwide Register of Laboratory Results for Research (RLRR),^25^ which includes results from biochemistry and haematology tests taken at hospitals and general practitioners in Denmark. We assessed the proportion of EBV capsid antigen IgM positive individuals who were also hospitalised with IM within 1 month and later went on to develop IBD to assess whether there was a difference in IBD development in those hospitalisation with laboratory confirmed IM compared to those not hospitalised with laboratory confirmed IM. All analyses were conducted in R (v4.3.0).

## RESULTS

We identified 40,244 individuals from the general Danish population between 1st January 1977 and 31st December 2021 with an IM diagnosis. 211 were excluded from inclusion in the severe IM arm based on a prevalent IBD diagnosis, with a further 349 excluded due to residence outside Denmark 10 years prior to IM diagnosis (see Figure 1 for flow diagram of study cohort inclusion). A total of 39,684 individuals were included in the severe IM cohort and matched to 396,840 IM control individuals without an IM diagnosis, based on sex, age, and residential municipality. These contributed a total 8,010,000 person-years of follow up to the analyses. There was a comparable proportion of males to females within the severe IM cohort (52.6% male) and most individuals were diagnosed with IM at 17-29 years of age (40.1%), only 4.7% of those with severe IM had a complication associated with their IM hospitalisation (including hepatosplenomegaly, neuropathy, and splenic rupture). Thirty-two percent of total severe IM cases were diagnosed between 2010-2021 (see Table 1 for baseline characteristics).

**Figure 1.**
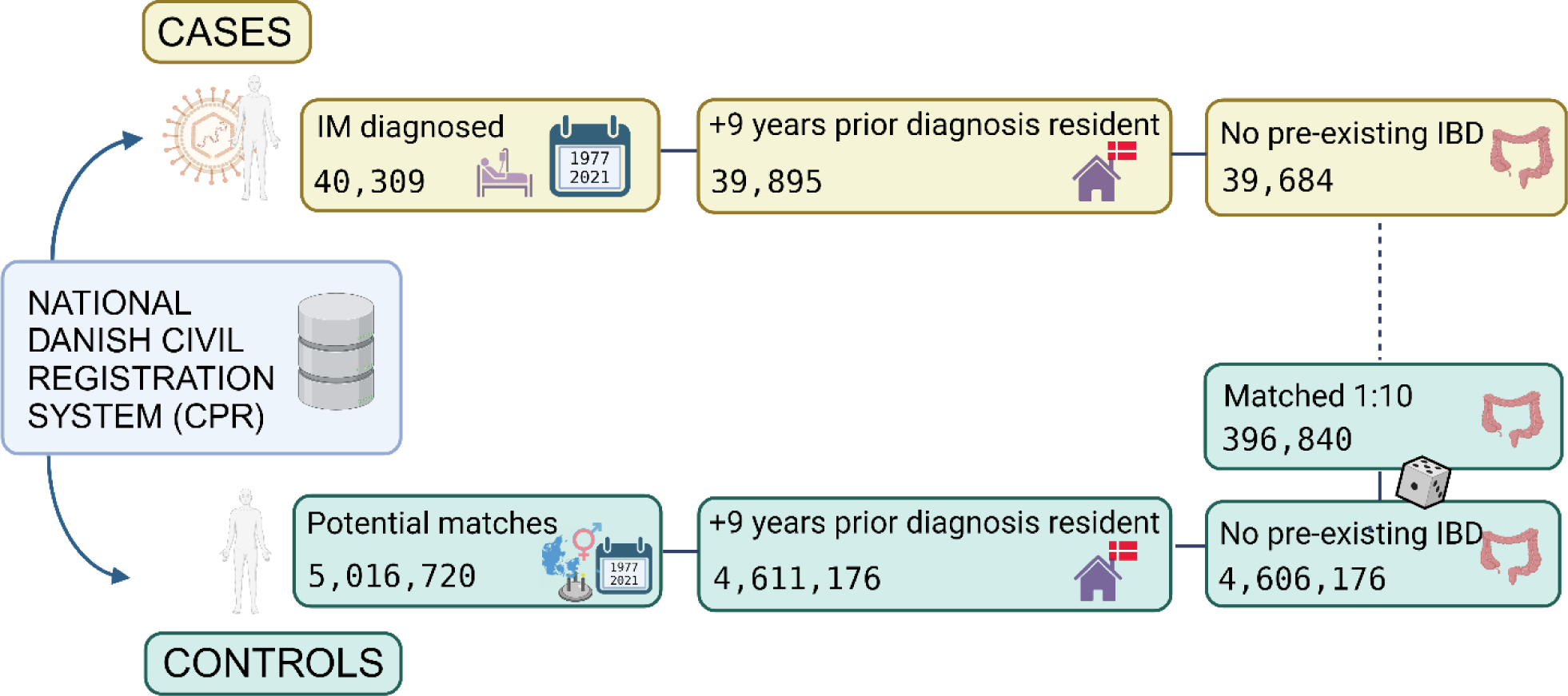
Cohort inclusion flow chart for severe IM and non-severe IM.

**Table 1.**
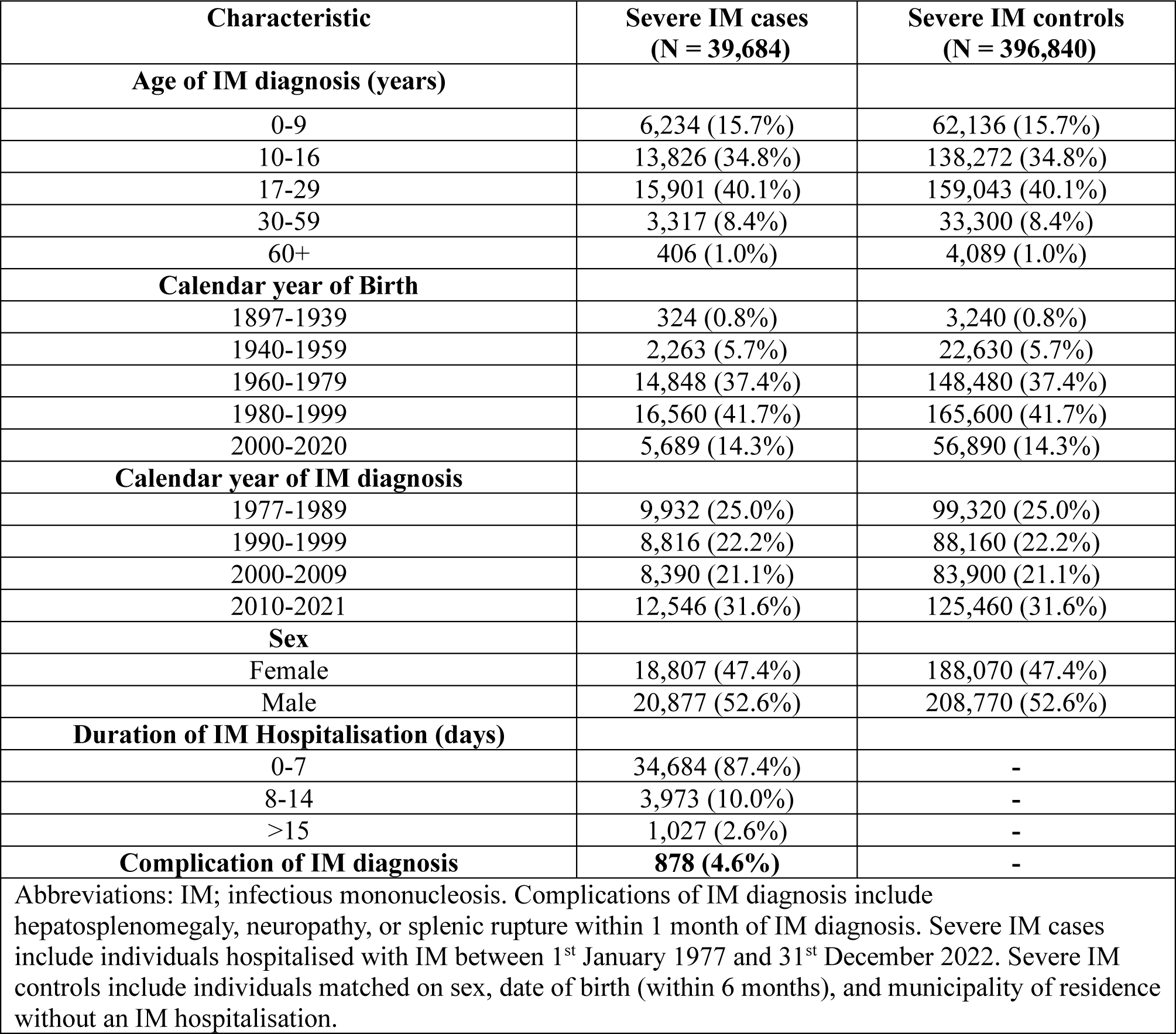
Baseline characteristics of severe and non-severe IM group at time of IM diagnosis.

| Characteristic | Severe IM cases<br>(N = 39,684) | Severe IM controls<br>(N = 396,840) |
| --- | --- | --- |
| **Age of IM diagnosis (years)** |  |  |
| 0-9 | 6,234 (15.7%) | 62,136 (15.7%) |
| 10-16 | 13,826 (34.8%) | 138,272 (34.8%) |
| 17-29 | 15,901 (40.1%) | 159,043 (40.1%) |
| 30-59 | 3,317 (8.4%) | 33,300 (8.4%) |
| 60+ | 406 (1.0%) | 4,089 (1.0%) |
| **Calendar year of Birth** |  |  |
| 1897-1939 | 324 (0.8%) | 3,240 (0.8%) |
| 1940-1959 | 2,263 (5.7%) | 22,630 (5.7%) |
| 1960-1979 | 14,848 (37.4%) | 148,480 (37.4%) |
| 1980-1999 | 16,560 (41.7%) | 165,600 (41.7%) |
| 2000-2020 | 5,689 (14.3%) | 56,890 (14.3%) |
| **Calendar year of IM diagnosis** |  |  |
| 1977-1989 | 9,932 (25.0%) | 99,320 (25.0%) |
| 1990-1999 | 8,816 (22.2%) | 88,160 (22.2%) |
| 2000-2009 | 8,390 (21.1%) | 83,900 (21.1%) |
| 2010-2021 | 12,546 (31.6%) | 125,460 (31.6%) |
| **Sex** |  |  |
| Female | 18,807 (47.4%) | 188,070 (47.4%) |
| Male | 20,877 (52.6%) | 208,770 (52.6%) |
| **Duration of IM Hospitalisation (days)** |  |  |
| 0-7 | 34,684 (87.4%) | - |
| 8-14 | 3,973 (10.0%) | - |
| >15 | 1,027 (2.6%) | - |
| **Complication of IM diagnosis** | **878 (4.6%)** | - |
| Abbreviations: IM; infectious mononucleosis. Complications of IM diagnosis include hepatosplenomegaly, neuropathy, or splenic rupture within 1 month of IM diagnosis. Severe IM cases include individuals hospitalised with IM between 1<sup>st</sup> January 1977 and 31<sup>st</sup> December 2022. Severe IM controls include individuals matched on sex, date of birth (within 6 months), and municipality of residence without an IM hospitalisation. |  |  |

We calculated an IBD incidence rate of 0.67 following severe IM group compared to 0.50 in those not hospitalised with IM with the median time to IBD diagnosis in the severe IM group being 19.3 years compared with 22.1 years in the control group (see Figure 2 for cumulative incidence of IBD, CD and UC). The crude HR for IBD diagnosis following severe IM was 1.35 (95% CI: 1.23-1.48) and this increased risk was seen in both CD and UC (see Table 2 for HR of IBD following severe IM). Following adjustment for sex, age and municipality of residence, this increased risk of IBD remained significantly increased (HR: 1.35; 95% CI: 1.22-1.49) and was again particularly seen in CD (HR: 1.56; 95% CI: 1.34-1.83) compared with UC (HR: 1.23; 95% CI: 1.08-1.40). This association was also found in both females (HR: 1.36; 95% CI:1.20-1.55) and males (HR: 1.34; 95% CI: 1.17-1.54) but only significantly associated with those diagnosed with IM at 10-16 years (HR:1.42; 95% CI:1.22-1.64) and 17-29 years (HR:1.34; 95% CI: 1.15-1.56). A significantly increased risk of IBD diagnosis following severe IM in 0-9 years was only observed in CD (HR: 1.77; 95% CI: 1.26-2.49). See Figures 3 and 4 for HR for CD and UC development following severe IM, respectively.

**Figure 2.**
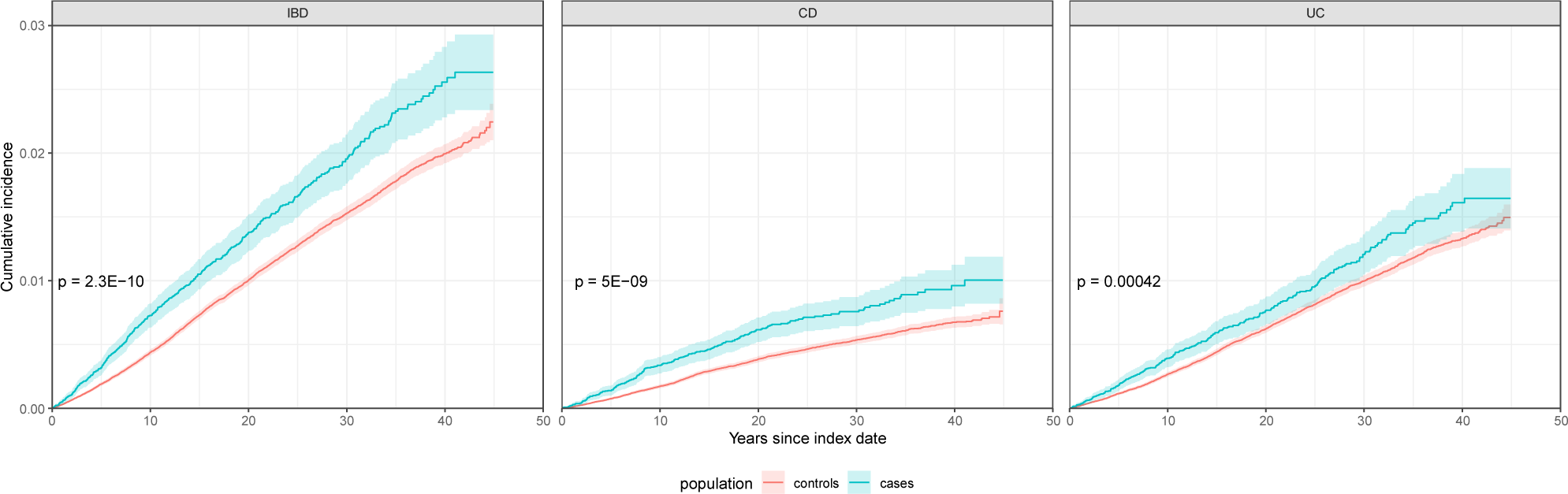
Cumulative incidence plots for IBD, CD and UC diagnosis in severe IM (cases) compared with non-severe IM (controls)

**Figure 3.**
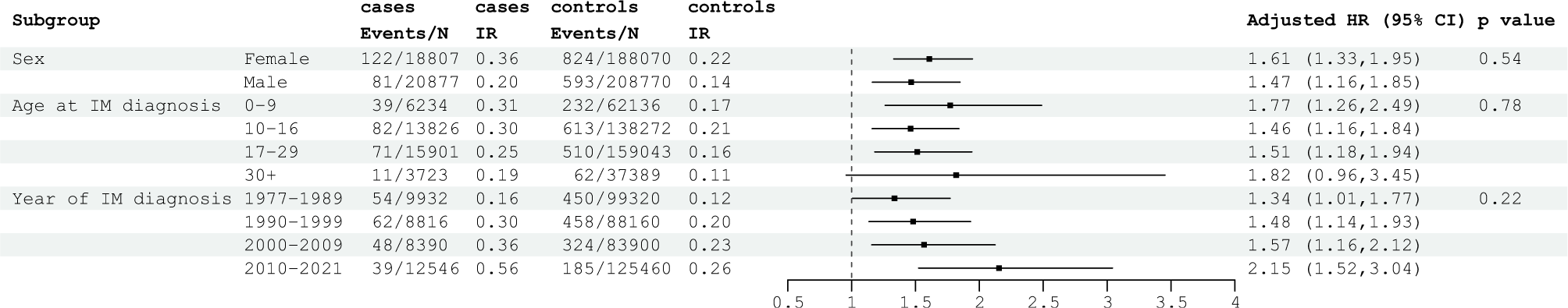
Crude incidence rates and adjusted hazard ratio for CD diagnosis in severe IM compared with non-severe IM stratified by sex, age at IM diagnosis and year of IM diagnosis.

**Figure 4.**
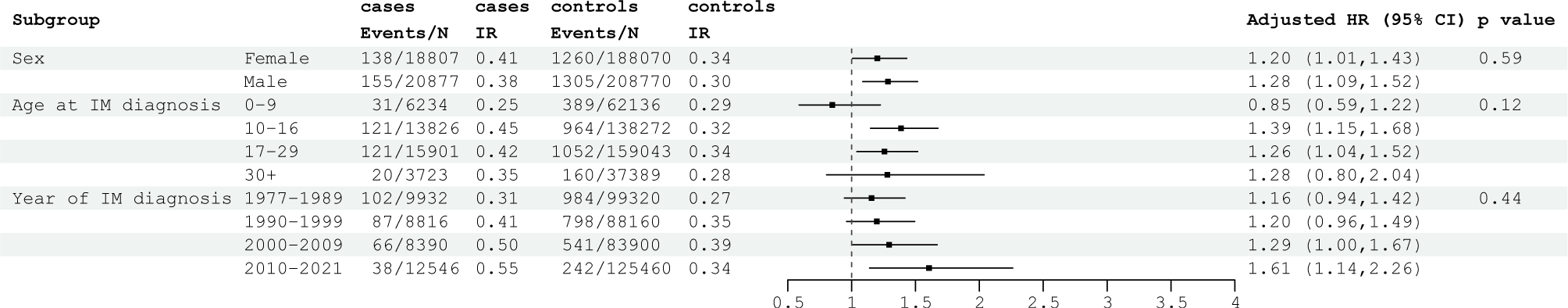
Crude incidence rates and adjusted hazard ratio for UC diagnosis in severe IM compared with non-severe IM stratified by sex, age at IM diagnosis and year of IM diagnosis.

**Table 2.** Crude incidence, crude HR, and adjusted HR for IBD, CD and UC diagnosis following severe IM compared with non-severe IM, stratified by sex, age at IM diagnosis and year of IM diagnosis.

|  | Severe IM cases<br>(N = 39,684) |  |  |  | Severe IM controls<br>(N = 396,840) |  |  |  | Crude HR | Crude HR 95% CI | Crude p-value | Adjusted HR | Adjusted* HR 95% CI | Adjusted* p-value |
| --- | --- | --- | --- | --- | --- | --- | --- | --- | --- | --- | --- | --- | --- | --- |
|  | Total follow-up (100,000 person-years) | IBD events in severe IM (N) | Total severe IM (N) | IBD IR (per 1000 person years) | Total follow-up (100,000 person-years) | IBD events in controls (N) | Total controls (N) | IBD IR (per 1000 person years) |  |  |  |  |  |  |
| **IBD** | 7.42 | 496 | 39,684 | 0.67 | 80.1 | 3982 | 396,840 | 0.5 | 1.35 | 1.23- 1.48 | <0.001 | 1.35 | 1.22-1.49 | <0.001 |
| **CD** | 7.42 | 203 | 39,684 | 0.27 | 80.1 | 1417 | 396,840 | 0.18 | 1.55 | 1.33-1.79 | <0.001 | 1.56 | 1.34-1.83 | <0.001 |
| **UC** | 7.42 | 293 | 39,684 | 0.40 | 80.1 | 2565 | 396,840 | 0.32 | 1.24 | 1.10-1.40 | <0.001 | 1.23 | 1.08-1.40 | 0.002 |
| **Sex** |  |  |  |  |  |  |  |  |  |  |  |  |  |  |
| Female | 3.38 | 260 | 18,807 | 0.77 | 36.8 | 2084 | 188,070 | 0.57 | 1.36 | 1.20-1.55 | 0.84 | 1.36 | 1.20-1.55 | 0.86 |
| Male | 4.03 | 236 | 20,877 | 0.58 | 43.3 | 1898 | 208,770 | 0.44 | 1.34 | 1.17-1.53 |  | 1.34 | 1.17-1.54 |  |
| **Age at IM diagnosis** |  |  |  |  |  |  |  |  |  |  |  |  |  |  |
| 0-9 | 1.26 | 70 | 6,234 | 0.55 | 13.4 | 621 | 62,136 | 0.46 | 1.20 | 0.94-1.53 | 0.69 | 1.20 | 0.93-1.53 | 0.69 |
| 10-16 | 2.71 | 203 | 13,826 | 0.75 | 29.8 | 1577 | 138,272 | 0.53 | 1.42 | 1.23-1.64 |  | 1.42 | 1.22-1.64 |  |
| 17-29 | 2.87 | 192 | 15,701 | 0.67 | 31.1 | 1562 | 159,043 | 183 | 1.34 | 1.15-1.55 |  | 1.34 | 1.15-1.56 |  |
| ≥30 | 0.57 | 31 | 3,723 | 0.55 | 5.8 | 222 | 37,389 | 140 | 1.43 | 0.98-2.08 |  | 1.43 | 0.98-2.08 |  |
| **Calendar year of IM diagnosis** |  |  |  |  |  |  |  |  |  |  |  |  |  |  |
| 1977-1989 | 3.29 | 156 | 39,684 | 0.47 | 36.3 | 1434 | 99,320 | 144 | 1.21 | 1.03-1.43 | 0.047 | 1.21 | 1.03-1.43 | 0.05 |
| 1990-1999 | 2.10 | 149 | 39,684 | 0.71 | 22.8 | 1256 | 88,160 | 201 | 1.30 | 1.09-1.54 |  | 1.30 | 1.10-1.54 |  |
| 2000-2009 | 1.33 | 114 | 39,684 | 0.86 | 14.0 | 865 | 83,900 | 226 | 1.39 | 1.15-1.69 |  | 1.40 | 1.15-1.70 |  |
| 2010-2021 | 0.69 | 77 | 39,684 | 1.11 | 7.1 | 427 | 125,460 | 220 | 1.84 | 1.45-2.35 |  | 1.84 | 1.45-2.35 |  |
| Abbreviations: IM; infectious mononucleosis, IR; incidence rate, HR; hazard ratio, 95% CI; 95% confidence interval. Severe IM cases include individuals hospitalised with IM between 1<sup>st</sup> January 1977 and 31<sup>st</sup> December 2021. Severe IM controls include individuals matched on sex, date of birth (within 6 months), and municipality of residence without an IM hospitalisation to severe IM cases. |  |  |  |  |  |  |  |  |  |  |  |  |  |  |
| *adjusted for sex, age and municipality of residence as appropriate. |  |  |  |  |  |  |  |  |  |  |  |  |  |  |

When assessing the risk of severe IBD disease development following IM diagnosis, compared with matched non-severe IM individuals, we found that the risk of any IBD-related hospitalisation, corticosteroid use, surgery, use of immunomodulators, or use of biologics was not significantly associated with severe-IM compared with non-severe IM (HR: 1.05; 95% CI:0.93-1.19). This was also the case in those diagnosed with both CD (HR: 1.14; 95% CI:0.98-1.32) and UC (HR: 0.96; 95% CI:0.84-1.11) see Table 3 for HR for severe IBD phenotype following severe IM. With regards to the individual outcomes, all were insignificant; comprising hospitalisation (HR: 1.21; 95% CI:0.94-1.55), corticosteroid use (HR: 0.84; 95% CI:0.71-1.00), surgery (HR: 1.08; 95% CI:0.9-1.31), immunomodulator use (HR: 1.16; 95% CI:0.97-1.39), and biologics use (HR: 0.95; 95% CI:0.79-1.13). See Table 3 for risk of severe IBD following severe IM.

**Table 3.** Crude incidence, crude HR, and adjusted HR for severe IBD composite outcome (including IBD related hospitalisation, corticosteroid use, surgery, immunomodulators or biologics use) following severe IM compared with non-severe IM.

|  | Severe IM cases<br>(N = 39,684) |  |  |  | Severe IM controls<br>(N = 396,840) |  |  |  | Crude<br>HR | Crude<br>HR 95%<br>CI | Adjusted*<br>HR | Adjusted*<br>HR 95%<br>CI |
| --- | --- | --- | --- | --- | --- | --- | --- | --- | --- | --- | --- | --- |
|  | Total<br>severe IM<br>follow-up<br>(person<br>years) | Severe<br>IBD<br>events in<br>severe IM<br>(N) | Total<br>IBD in<br>severe<br>IM (N) | IBD IR<br>in<br>severe<br>IM | Total<br>controls<br>follow-up<br>(person<br>years) | Severe<br>IBD<br>events<br>controls<br>(N) | Total IBD<br>in<br>controls<br>(N) | IBD IR<br>in<br>controls |  |  |  |  |
| **IBD** | 1489 | 275 | 390 | 185 | 13002 | 2127 | 3150 | 164 | 1.09 | 0.96-1.23 | 1.05 | 0.93-1.19 |
| **CD** | 296 | 136 | 156 | 459 | 2527 | 871 | 1071 | 345 | 1.20 | 1.00-1.43 | 1.14 | 0.98-1.32 |
| **UC** | 1193 | 139 | 234 | 117 | 10475 | 1256 | 2079 | 120 | 0.97 | 0.81-1.15 | 0.96 | 0.84-1.11 |
| **hospitalisation** | 2137 | 195 | 390 | 91 | 19262 | 1426 | 3150 | 74 | 1.16 | 1.00-1.35 | 1.21 | 0.94-1.55 |
| **corticosteroid** | 2368 | 217 | 390 | 92 | 18326 | 1704 | 3150 | 93 | 1.01 | 0.87-1.16 | 0.84 | 0.71-1.00 |
| **surgery** | 3688 | 70 | 390 | 19 | 30910 | 472 | 3150 | 15 | 1.22 | 0.95-1.57 | 1.08 | 0.90-1.31 |
| **immunomodulators** | 3134 | 140 | 390 | 45 | 24154 | 1246 | 3150 | 52 | 0.90 | 0.75-1.07 | 1.16 | 0.97-1.39 |
| **biologics** | 3687 | 126 | 390 | 34 | 30545 | 884 | 3150 | 29 | 1.17 | 0.97-1.41 | 0.95 | 0.79-1.13 |

Abbreviations: IM; infectious mononucleosis, IR; incidence rate, HR; hazard ratio, 95% CI; 95% confidence interval. Severe IM cases include individuals hospitalised with IM between 1<sup>st</sup> January 1977 and 31<sup>st</sup> December 2021. Severe IM controls include individuals matched on sex, date of birth (within 6 months), and municipality of residence without an IM hospitalisation to severe IM cases.
\*adjusted for sex, age and municipality of residence.

We identified seventy-three IM negative individuals with a chlamydia trachomatis diagnosis for negative-control matching and found that severe IM patients were not at significantly increased risk of IBD development compared to chlamydia patients (HR: 1.24; 95% CI: 0.93-1.65). Finally, we used the RLRR to assess whether there may be a difference in IBD diagnosis in confirmed EBV positive infectious mononucleosis cases that were hospitalised (severe IM), compared to confirmed EBV positive infectious mononucleosis cases who were not hospitalised by identifying positive test results for EBV capsid antigen IgM. We limited analysis to samples taken at Køge hospital to ensure complete test coverage for a given population area and found with 3864 individuals with a positive test who later go on to develop IBD. Of these, 460 were hospitalised with IM around the time of testing positive (+/-31 days) compared with 3404 with a positive test who were not hospitalised, giving a hospitalisation fraction of 0.11 of potential infectious mononucleosis cases in the area. Survival analysis showed an OR 1.7 (95% CI: 057-4.99: p=0.344) for IBD development following hospitalisation with a positive test compared to not being hospitalised with a positive test.

## DISCUSSION

In this population-representative, nationwide cohort study we follow almost 40,000 patients hospitalised with IM and over 396,000 sex-, age-, and municipality-matched control individuals for the development of IBD for over 8 million person-years to identify a 35% increased risk of IBD development following severe IM. This was seen particularly for CD (56% increased risk) but also present for UC (23% increased risk). There was no difference in risk based on sex, age at IM diagnosis, or calendar year of IM diagnosis. We do not identify a significant association between hospitalisation with IM and later development of severe IBD (including IBD-related hospitalisation, surgery, or treatment escalation).

The main findings of this study indicate that hospitalisation with IM is associated with a significantly increased risk of IBD development. These findings are in keeping with those recently published by Loosen et al.,^24^who identify an increased risk of IBD following primary care diagnosis with IM in a subset of German practices at the same rate as that of this current study (HR:1.35; 95% CI: 1.01-1.81).^24^ Although no other cohort studies to date have explored the association between infectious mononucleosis and IBD development, the consistent findings between these two large cohorts indicates a possible underlying pathophysiological contribution of IM disease to IBD development.

Most infectious mononucleosis cases are caused by the common and highly transmissible gamma herpes virus, EBV, with seroconversion occurring at the highest rates in early childhood and adolescence.^14^ This high prevalence of seropositivity at an early period in life makes identifying putative associations between EBV and subsequent IBD development highly challenging, hence the focus of this study on severe IM or a subset of the severe clinical manifestation of EBV infection. To overcome the challenge of high EBV seroprevalence in detecting association with IBD Nandy et. al.,^23^ have recently attempted to apply unbiased, comprehensive serological viral screening to a small longitudinal cohort of 42 IBD patients with pre- and post-diagnostic serum samples. Authors show that EBV exposure levels are significantly associated with CD patients compared with healthy controls (CD=78.2% vs healthy controls=53.3%, p=0.045), suggesting a pathogenic role for EBV in either the onset or progression of CD. Furthermore, longitudinal analysis on the same cohort indicated this association may be putative as increased levels of EBV exposure were present as early as 5 years before disease diagnosis.^23^

The vast majority of individuals are infected with EBV during childhood or adolescence, resulting in a persistent and mostly latent EBV infection, which most commonly manifests as IM, when primary infection occurs in adolescence.^16^ A Danish study assessing age-dependent variations both in IM attack rates and EBV seroconversion hazard rates found 85% of the Danish population had seroconverted by age 30 years,^15^ with peak incidence being in the 15-24 year age range. This is mirrored in the age distribution of the IM cases identified in the current study, where we find a corresponding stronger effect sizes for IBD development seen in those diagnosed with IM in 10-16- and 17-29-year age range for both CD and UC. Interestingly, unlike Loosen et al., who identify this association between IM and later IBD development only for CD subtype, we also find this increased risk is significantly associated with UC development. The presence of EBV-induced gene 3 and EBV-encoded small RNA1, indicative of activity of EBV-infected cells, has previously been associated with both CD and UC, particularly during periods of inflammatory disease activity.^27,28^ Additionally, recent exploration of gene regulatory regions and transcription factors using GWAS catalogues for multiple autoimmune diseases identify particularly strong associations with EBV nuclear antigen 2 (EBNA2), an EBV derived transcription factor, and ulcerative colitis (p<10^-8^). Supporting the potential role of genetic mechanisms dependent on EBNA2 in the induction of disease in individuals susceptible to ulcerative colitis.^29^

Although linked to several cancers, including Hodgkin’s Lymphoma, a high proportion of Burkitt’s lymphomas, several nasopharyngeal carcinomas,^30^ and lymphoproliferative disorders associated with transplantation and immunosuppression,^31^ it is only in recent years that EBV and IM have been implicated in the development of complex, immune-mediated diseases.^32–34^ Numerous mechanisms have been hypothesised for the potential contribution of EBV in the development of these diseases. Most recently, clonally expanded B-lymphocytes have been demonstrated to be cross-reactive for the central nervous system protein glial cell adhesion molecule and EBNA1, exposing their high-affinity molecular mimicry in multiple sclerosis.^35,36^ Similarly, cross-reactive molecularly mimicry may be at play in the immune tissue of the gastrointestinal mucosa or against gastrointestinal commensals in IBD. ^37,38^ Alternatively, under the immune exhaustion hypothesis, EBV infection in the acute phase could be exhausting the immune system’s resources, potentially increasing vulnerability to IBD in genetically susceptible individuals as is also seen in MS with EBV-specific T-lymphocytes eventually becoming exhausted through cytokine polyfunctionality.^39^

Infectious mononucleosis is characterised by an aggressive peripheral CD8^+^ T cell lymphocytosis, that gives the disease its name. It is likely that it is the extent of the expansion of these activated T-cells that contributes to the clinical features of IM, and the magnitude of this response is the differentiating factor in those developing severe IM and being hospitalised. Individuals expressing certain HLA alleles, including HLA-A2 and HLA-B8 are more likely to express high levels of circulating CD8^+^ T cells against EBV lytic proteins.^40^ We know that HLA region plays a key role in IBD disease pathophysiology, with this genetic region being where the most significant risk variants and the largest effect sizes for IBD susceptibility are located.^41^ It is plausible therefore, that HLA class I gene region alleles with HLA traits A and B implicated in IBD development might also be playing a role in the adaptive immune response to EBV and severe IM development.

A particular strength of this current study lies in the use of highly reliable population-representative, nationwide health registry data ensuring that all diagnoses of both infectious mononucleosis and inflammatory bowel disease captured are reliable. Danish registries allow for complete follow-up due to complete data on emigration and death, with no loss to follow-up when moving within the country. Due to nationwide registration that is used for state reimbursement, there is also a low risk of misclassification of outcomes. Our sensitivity analysis evaluating whether risk of any EBV positive test was associated with IBD development did not show a significant difference in odds for those hospitalised with IM and the hospitalisation fraction identified in this current study (0.1) is similar to that found in previous Danish data (0.07), indicating the generalisability and replicability of our findings.^15^ Additionally, using these data, we were able to identify infectious mononucleosis patients from 1977-2021, providing 44 years of follow up, which is reflected in the size of the cohort included in our analyses. This longer duration of follow-up is particularly important in the exploration of associations between early life events and later, chronic disease development, as is the case for the current study. Of particular importance in this study is our use of severe IM (i.e., hospitalisation with infectious mononucleosis) as an exposure, which has not been previously done and allows for the use of a consistent and biologically plausible marker of innate immune dysfunction following EBV infection in the development of IBD. Finally, an important strength of this work is the use of Danish IBD diagnoses which have previously been validated and found to be close to complete (95%) and highly valid (90-97%) when using 2 diagnoses for ascertainment as is the case for the current study.^42,43^

Whilst we use high-quality data with reliable diagnoses of both infectious mononucleosis and IBD, there are some limitations to the current study. The observational design has within it an inherent risk of bias due to the inability to control for unknown or unmeasured confounders such as health seeking behaviours in the cohort. Although, this may have impacted on our findings of a difference in the risk of later IBD diagnosis in the severe IM group (hospitalised with IM) compared with control group (not hospitalised with IM), it is unlikely to have resulted in a systemic bias that could result in the large effect size and strong significance detected. Reassuringly, our null finding of increased IBD-related hospitalisation in severe IM compared with controls, is not only an indication of similar disease severity following IM hospitalisation but also innately indicative of similar health seeking behaviours around the time of and following IBD diagnosis. Another limitation is the lack of data on smoking habits which may be a potential confounder as recent data has indicated an interaction between current smoking status and infectious mononucleosis risk^44^ however if this were systemically biasing findings, we would not expect to see significant risk of UC diagnosis following severe IM, due to the protective nature of smoking in UC disease development and course.^45^

In conclusion, we identify an interesting association between severe infectious mononucleosis disease and subsequent development of inflammatory bowel disease, particularly Crohn’s disease. Further exploration of the potential role of EBV infection in IBD pathophysiology is warranted.

## Data Availability

The study was based on data from the Danish National Health registers (https://sundhedsdatastyrelsen.dk). The register
data are protected by the Danish Act on Processing of Personal Data and are accessed through application to and approval from the Danish Data Protection Agency and the Danish Health Data Authority.

## ACKNOWLEDGEMENTS

With thanks to Manasi Agrawal, Heidi Søgaard Christensen, and Rasmus Froberg Brøndum for their contribution to the earlier work undertaken exploring the EBV testing using the Danish nationwide Register of Laboratory Results for Research and Jesus Vicente Torresano Lominchar for this contribution in the conception and creation of some manuscript tables and figures.

## SUPPLEMENTARY MATERIAL

**Supplementary Table 1.** List of diagnostic codes.

| DEFINITION | CODES |
| --- | --- |
| **Infectious mononucleosis** | ICD-8: 075, ICD-10: B27 |
| **Complications of infectious mononucleosis** | Enlarged lymph nodes (ICD-8: 782.79; ICD-10: R59)<br>Acute lymphadenitis (ICD-8: 683.99; ICD-10: L04)<br>Lymphadenitis (ICD-8: 289.29, 289.39; ICD-10: I88.0 I88.8)<br>Hepato-splenomegaly (ICD-8: 785.19, 782.89; ICD-10: R16)<br>Splenic rupture (ICD-8: 289.41, 289.43; ICD-10: D73.5) |
| **Inflammatory bowel disease** | Crohn's disease (ICD-8: 563.01, 563.02, 563.08, 563.09; ICD-10: K50)<br>Ulcerative colitis (ICD-8: 563.19, 569.04; ICD-10: K51) |
| **Chlamydia** | Chlamydia (ICD-8: 079.84; ICD-10: A56) |

**Supplementary Table 2.**
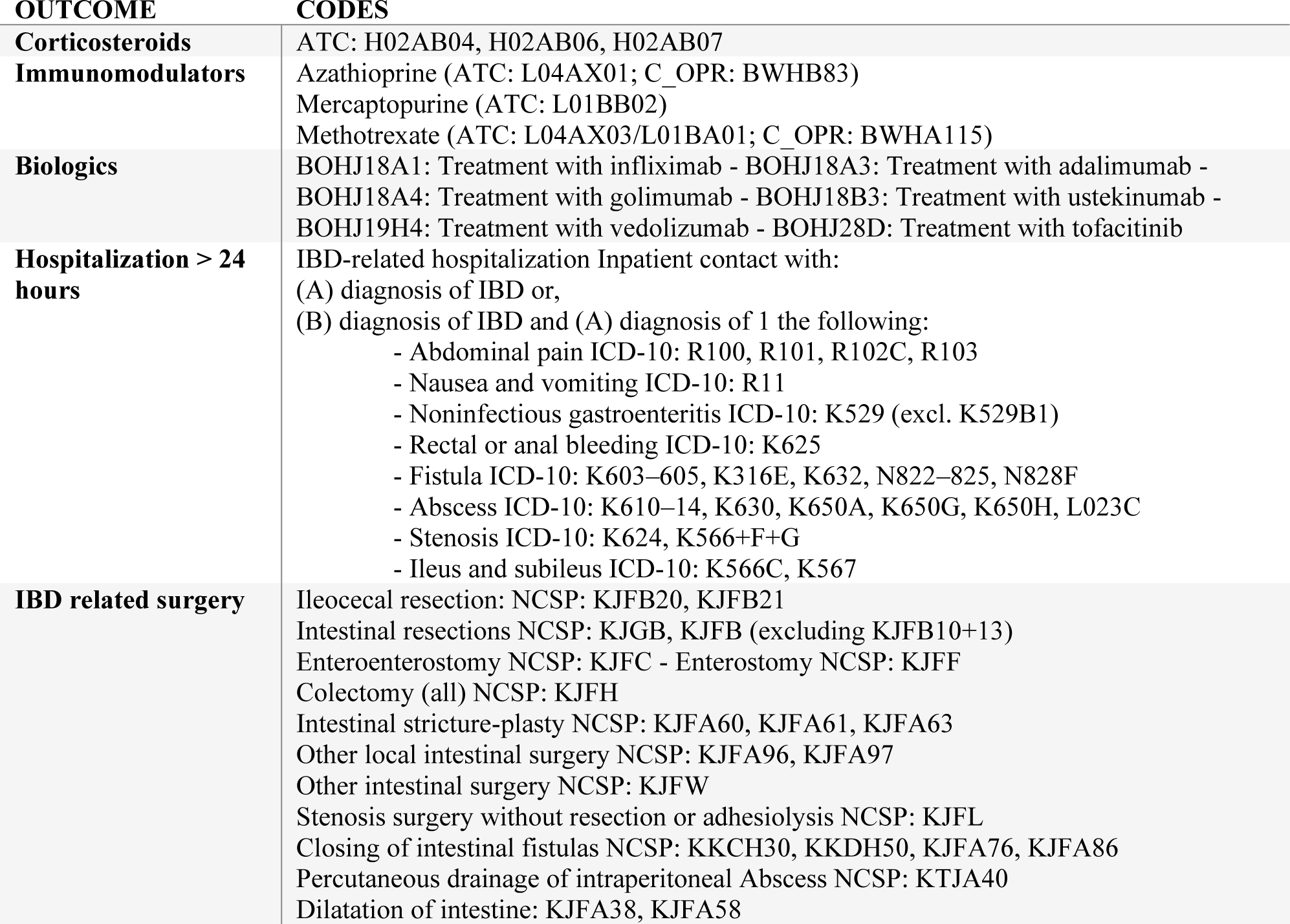
Definitions and associated diagnostic and procedure codes for outcomes associated with IBD disease progression.

